# Rapid growth in disposable e-cigarette vaping among young adults in Great Britain from 2021 to 2022: a repeat cross-sectional survey

**DOI:** 10.1101/2022.05.06.22274762

**Authors:** Harry Tattan-Birch, Sarah E Jackson, Loren Kock, Martin Dockrell, Jamie Brown

**Author notes:** **Correspondence to:** Harry Tattan-Birch, |, Address: Institute of Epidemiology and Health Care, 1-19 Torrington Place, Fitzrovia, London WC1E 7HB.

## Abstract

**Aims:** To estimate recent trends in the prevalence of disposable e-cigarette vaping in Great Britain, overall and across ages.

**Design:** The Smoking Toolkit Study, a monthly representative cross-sectional survey. Setting: Great Britain.

**Participants:** 29,976 adults (≥18 years) completed telephone interviews from January 2021 to January 2022.

**Measurements:** Current e-cigarette vapers were asked which type of device they mainly use. We estimated age-specific time trends in the prevalence of current disposable e-cigarette use among vapers and inhaled nicotine use (vaping/smoking) among adults.

**Findings:** From January 2021 to January 2022, there was a 14-fold increase in the percentage of vapers that used disposables, rising from 1.2% to 16.7% (prevalence ratio [PR], 14.4; 95%CI, 6.6-49.0). Growth in disposable e-cigarette vaping was most pronounced in younger adults (interaction p-value, .0.007): for example, the percentage of 18-year-old vapers using of disposables rose from 0.89% to 56.7% (PR, 64; 95%CI, 15-5790) while it rose from 1.3% to 6.2% (PR, 4.7; 95%CI, 1.7-144) among 45-year-old vapers. However, the overall percentage people currently using any inhaled nicotine, vaped or smoked, remained stable over time both among all adults (20.1% vs. 20.6%; PR, 1.03; 95%CI, 0.88-1.20) and among 18-year-olds (29.6% vs. 29.6%; PR, 1.00; 95%CI, 0.79-1.26).

**Conclusions:** Use of disposable e-cigarettes in Great Britain grew rapidly between 2021 and 2022, especially among younger adults, but the overall prevalence of inhaled nicotine use — smoked or vaped — was stable over time. Most young adult vapers in Great Britain now use disposable products.

## Introduction

Early electronic cigarettes (“e-cigarettes”) were disposable products that were poor at delivering nicotine. Over time, new e-cigarette types were developed to deliver nicotine contained in e-liquid more effectively through rechargeable devices with refillable tanks or replaceable pods (e.g. Juul).^1^ These devices came to dominate the global e-cigarette market and, by 2019, fewer than one-in-ten vapers used disposables in England or the US.^1–3^ Recently, a new form of disposable e-cigarette has started being sold under brand names like “Puff-bar”, “Elf-bar”, or “Geek-bar”.^4^ Unlike earlier disposables, these products deliver nicotine effectively using a similar technology to pod devices, including high-strength (20mg/ml in UK/EU) nicotine salts e-liquid.^5^ They retail for around £5 to £7 ($7 to$9) in the UK — about half the price of a pack of 20 cigarettes. US data show that, in 2021, disposables surpassed pods as the most commonly used type of e-cigarette among adolescents.^2^ Little is known about the popularity of disposables in other countries and older age groups. This study aims to estimate recent trends in the prevalence of disposable e-cigarette use in Great Britain, overall and across ages.

## Methods

### Design

The Smoking Toolkit Study (STS) is a monthly cross-sectional survey that recruits a nationally representative sample of adults (≥18 years) in Great Britain. It uses a hybrid of population and quota sampling. Great Britain is divided into areas of approximately 300 households, which are stratified by region and demographic profile before being selected at random to be included on the interview list. In selected areas, interviews are performed with one individual per household until age, employment status, and gender quotas are met. Raking is used to construct survey weights, adjusting data so that the demographic profile of the weighted sample matches that of Great Britain. This demographic profile is ascertained monthly using data from three sources: the 2011 UK Census, the Office for National Statistics mid-year estimates, and the annual National Readership Survey. Methods are described in detail elsewhere.^6^

### Participants

Participants completed telephone interviews between January 2021 and January 2022, inclusive. University College London Ethics Committee provided approval for the study (0498/001), and participants gave oral informed consent. All methods were carried out in accordance with relevant regulations.

### Measures

Smoking status was ascertained by asking participants which of the following applies to them: i) “I smoke cigarettes (including hand-rolled) every day”, ii) “I smoke cigarettes (including hand-rolled), but not every day”, iii) “I do not smoke cigarettes at all, but I do smoke tobacco of some kind (eg. Pipe, cigar or shisha)”, iv) “I have stopped smoking completely in the last year”, v) “I stopped smoking completely more than a year ago”, vi) “I have never been a smoker (i.e. smoked for a year or more)”. Participants were told that this question referred to cigarettes and other kinds of tobacco, not e-cigarettes or heat-not-burn products. Participants selecting i to iii were classified current smokers, iv and v former smokers, and vi never smokers.

Participants were asked whether they were currently using e-cigarettes to cut down on the amount they smoke, in situations when they are not allowed to smoke, to help them stop smoking, or for any other reason. Those who responded positively to any of these questions were considered current vapers.

Current vapers were asked which type of device they mainly use. Those who responded, “a disposable e-cigarette or vaping device (non-rechargeable)”, were considered disposable e-cigarette vapers.

### Analysis

Weighted logistic regression was used to estimate time trends in the proportion of (i) adults and (ii) current vapers who use disposable e-cigarettes, overall and after including time, age and their interaction as predictors. Time and age were modelled using restricted cubic splines, with estimates for 18-, 25-, 35- and 45-year-olds reported to illustrate how trends differed across ages. We ran analogous analyses to estimate time trends in the proportion of adults who currently (i) vape, (ii) smoke, or (iii) use any form of inhaled nicotine — be that smoked or vaped. Note that prevalence of disposable e-cigarette use was very low in older age groups, which meant we were unable to estimate time trends in these groups. Finally, we reported the percentage of disposable e-cigarette vapers who reported being current, former, or never smokers. R version 4.1.0 was used for analyses (code: https://osf.io/km3x6/). Results were reported alongside 95% compatibility intervals (95%CIs).^7,8^

### Role of the funding source

The funders of the Smoking Toolkit Study had no role in the study design or conduct; collection, management, analysis, or interpretation of data; preparation, review or approval of the manuscript; or the decision to submit the manuscript for publication.

## Results

Of the 29,976 adults interviewed, 51.4% were women, and the average age was 52.6 years (SD, 18.6). From January 2021 to January 2022, there was a 14-fold increase in the percentage of vapers that used disposables, rising from 1.2% to 16.7% (prevalence ratio [PR], 14.4; 95%CI, 6.6-49.0). Overall, the prevalence of disposable e-cigarette use increased from 0.08% to 1.35% (Table 1; PR, 16.2; 95%CI, 5.8-45.1).

**Table 1:** Age-specific trends in current vaping, smoking and disposable e-cigarette vaping prevalence in Great Britain. Weighted prevalence estimates from logistic regression allowing an interaction between age and month, modelled non-linearly using restricted cubic splines (three knots). Data, analysis code, and estimates for other months available online (https://osf.io/km3x6/).

|  | Prevalence |  | Prevalence Ratio (95% CI) |
| --- | --- | --- | --- |
|  | Jan-21 | Jan-22 |  |
| Currently use inhaled nicotine (vaped or smoked) |  |  |  |
| 18-year-olds | 29.6% | 29.6% | 1.00 (0.79-1.26) |
| 25-year-olds | 28.6% | 30.1% | 1.05 (0.93-1.20) |
| 35-year-olds | 26.1% | 28.3% | 1.09 (0.97-1.22) |
| 45-year-olds | 21.9% | 23.4% | 1.07 (0.95-1.21) |
| All adults | 20.1% | 20.6% | 1.03 (0.88-1.20) |
| Currently vaping |  |  |  |
| 18-year-olds | 11.3% | 14.6% | 1.29 (0.84-2.00) |
| 25-year-olds | 10.8% | 13.7% | 1.27 (1.01-1.63) |
| 35-year-olds | 9.5% | 11.5% | 1.21 (0.99-1.49) |
| 45-year-olds | 7.7% | 8.6% | 1.12 (0.90-1.38) |
| All adults | 7.1% | 8.0% | 1.13 (0.97-1.31) |
| Currently smoking |  |  |  |
| 18-year-olds | 24.0% | 20.4% | 0.85 (0.62-1.11) |
| 25-year-olds | 22.6% | 20.5% | 0.91 (0.77-1.06) |
| 35-year-olds | 19.9% | 19.2% | 0.97 (0.85-1.10) |
| 45-year-olds | 16.3% | 16.0% | 0.98 (0.85-1.13) |
| All adults | 15.1% | 14.3% | 0.94 (0.85-1.05) |
| Currently vaping disposables |  |  |  |
| 18-year-olds | 0.08% | 9.69% | 115 (30-7310) |
| 25-year-olds | 0.11% | 3.50% | 31.3 (11.0-232) |
| 35-year-olds | 0.13% | 1.06% | 8.21 (2.12-40.9) |
| 45-year-olds | 0.10% | 0.53% | 5.18 (1.92-67.9) |
| All adults | 0.08% | 1.35% | 16.2 (5.82-45.1) |

Growth in disposable e-cigarette vaping was most pronounced in the youngest participants (Figure 1; interaction p-value, .0066). For instance, prevalence of disposable use among 45-year-old vapers rose from 1.3% to 6.2% (PR, 4.7; 95%CI, 1.7-144), whereas among 18-year-old vapers it increased from 0.89% to 56.7% (PR, 64; 95%CI, 15-5790).

**Figure 1.**
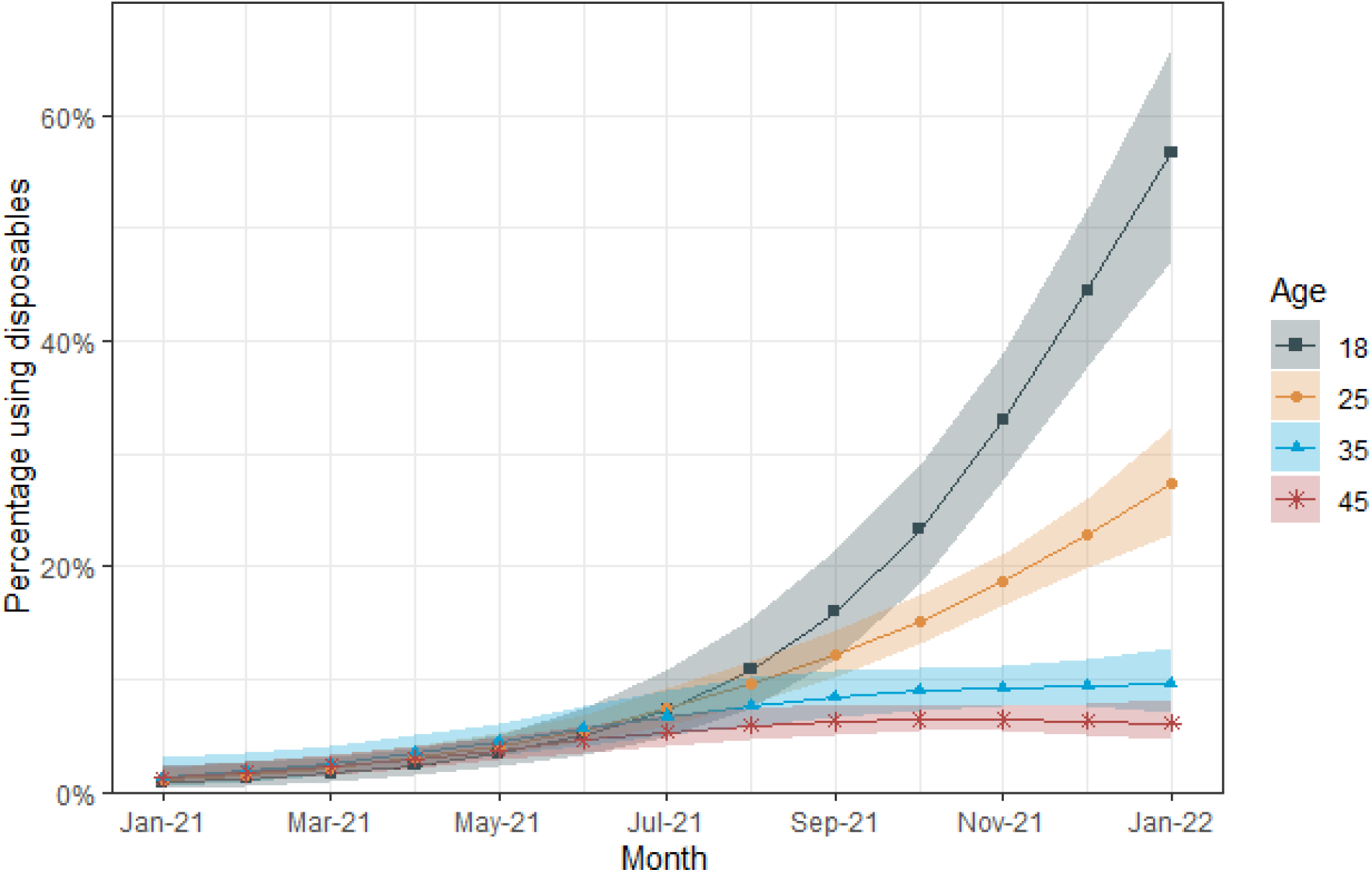
Percentage of current vapers using disposable e-cigarettes across ages in Great Britain from 2021 to January 2022. A total of 29,976 adults were surveyed (approximately 2,300 each month). Lines represent point estimates from logistic regression allowing an interaction between age and month, modelled non-linearly using restricted cubic splines (three knots). Shaded areas represent standard errors. Data and analysis code available online (https://osf.io/km3x6/)

Despite this, the overall percentage of adults currently using any inhaled nicotine (smoked or vaped) was stable over the study period (Table 1; 20.1% vs. 20.6%; PR, 1.03; 95%CI, 0.88-1.20). Even among young adults, where the rise in disposable vaping was most pronounced, inhaled nicotine use did not increase over time, estimated to be 29.6% for 18-year-olds in both January 2021 and January 2022 (Table 1; PR, 1.00; 95%CI, 0.79-1.26). Supplementary figures 1 to 3 show monthly trends in the prevalence of inhaled nicotine use, vaping, and smoking among adults of different ages.

Most disposable e-cigarette vapers were current (72.8%) or former smokers (18.4%), with few reporting never having smoked regularly (8.8%). Moreover, the proportion of disposable vapers who also smoked was similar across ages and over time (Supplementary figures 4 and 5).

## Discussion

Use of disposable e-cigarettes rose sharply between 2021 and 2022 in Great Britain — with the most rapid growth observed among the youngest adults, mirroring trends observed in US adolescents.^2^ At the start of 2021, fewer than 1% of 18-year-old vapers used disposables. This increased substantially throughout the past year, such that now over half of 18-year-old vapers report mostly using disposables. Despite this, the overall percentage of young people using any form of inhaled nicotine — be that smoked or vaped — was stable over time. This suggests that, in Great Britain up to 2022, disposable e-cigarettes have primarily attracted those who would otherwise use rechargeable devices or cigarettes, rather than those who would otherwise not use any nicotine product. Nonetheless, patterns of nicotine product use can change rapidly. Early and routine publication of data such as these are needed to guide policy and research. Study limitations include the wide 95%CIs around prevalence ratios due to few participants reporting disposable e-cigarette use in early months. Future studies should examine why disposable e-cigarettes have become the product of choice among young people in Great Britain and the US,^2^ and whether similar trends have occurred in other countries.

## Data Availability

Data and analysis code are available online

https://osf.io/km3x6/

## Conflict of Interest Disclosures

HTB, LK, MD, and SJ declare no conflicts of interest. JB has received unrestricted research funding to study smoking cessation from manufacturers of smoking cessation medications (Pfizer and Johnson & Johnson).

## Ethical Approval

University College London Ethics Committee provided approval for the study (0498/001), and participants gave oral informed consent.

## Funding

Cancer Research UK provides funding for the Smoking Toolkit Study in England and salary support for SJ (PRCRPG-Nov21\100002). The UK Prevention Research Partnership (MR/S037519/1) funds the Smoking Toolkit Study in Scotland and Wales and provides salary support for LK. The UK Prevention Research Partnership is funded by the British Heart Foundation, Cancer Research UK, Chief Scientist Office of the Scottish Government Health and Social Care Directorates, Engineering and Physical Sciences Research Council, Economic and Social Research Council, Health and Social Care Research and Development Division (Welsh Government), Medical Research Council, National Institute for Health Research, Natural Environment Research Council, Public Health Agency (Northern Ireland), The Health Foundation and Wellcome. HTB’s studentship is funded by the Office for Health Improvement and Disparities, previously Public Health England (558585/180737).

---

**Supplementary Figure 1.**
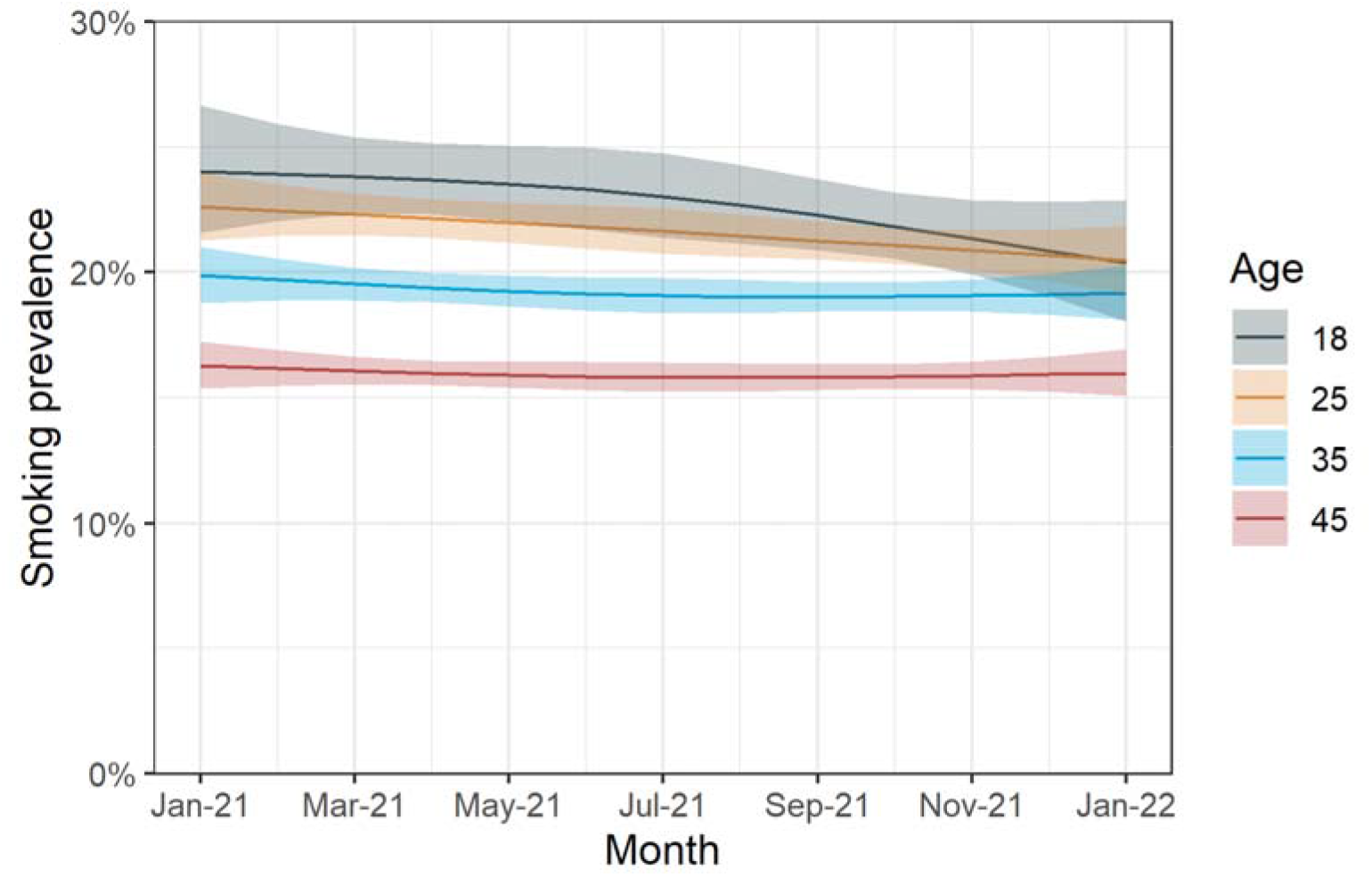
Smoking prevalence across ages in Great Britain from 2021 to January 2022. A total of 29,976 adults were surveyed (approximately 2,300 each month). Lines represent point estimates from logistic regression allowing an interaction between age and month, modelled non-linearly using restricted cubic splines. Shaded areas represent standard errors.

**Supplementary Figure 2.**
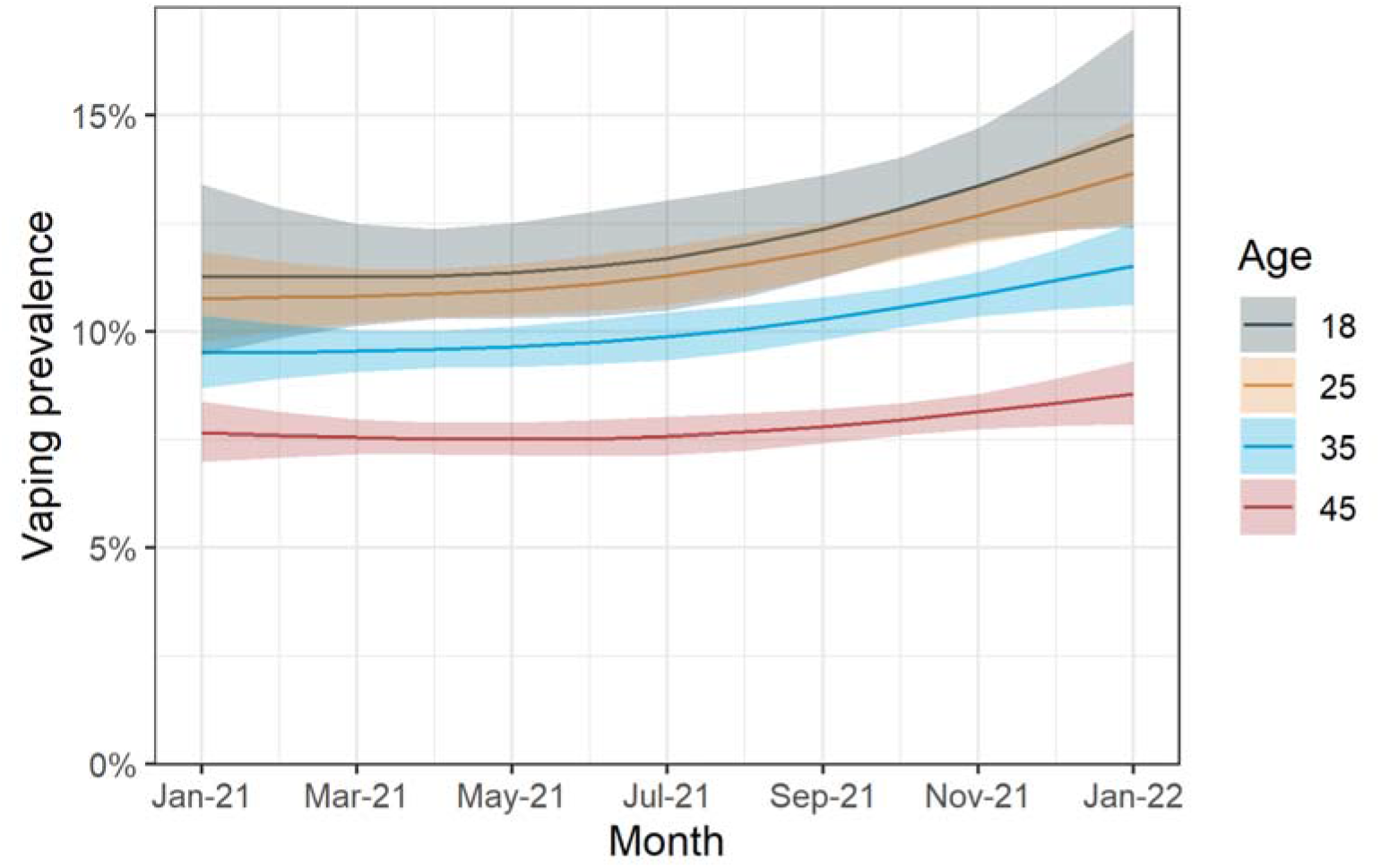
Vaping prevalence across ages in Great Britain from 2021 to January 2022. A total of 29976 adults were surveyed (approximately 2300 each month). Lines represent point estimates from logistic regression allowing an interaction between age and month, modelled non-linearly using restricted cubic splines. Shaded areas represent standard errors.

**Supplementary Figure 3.**
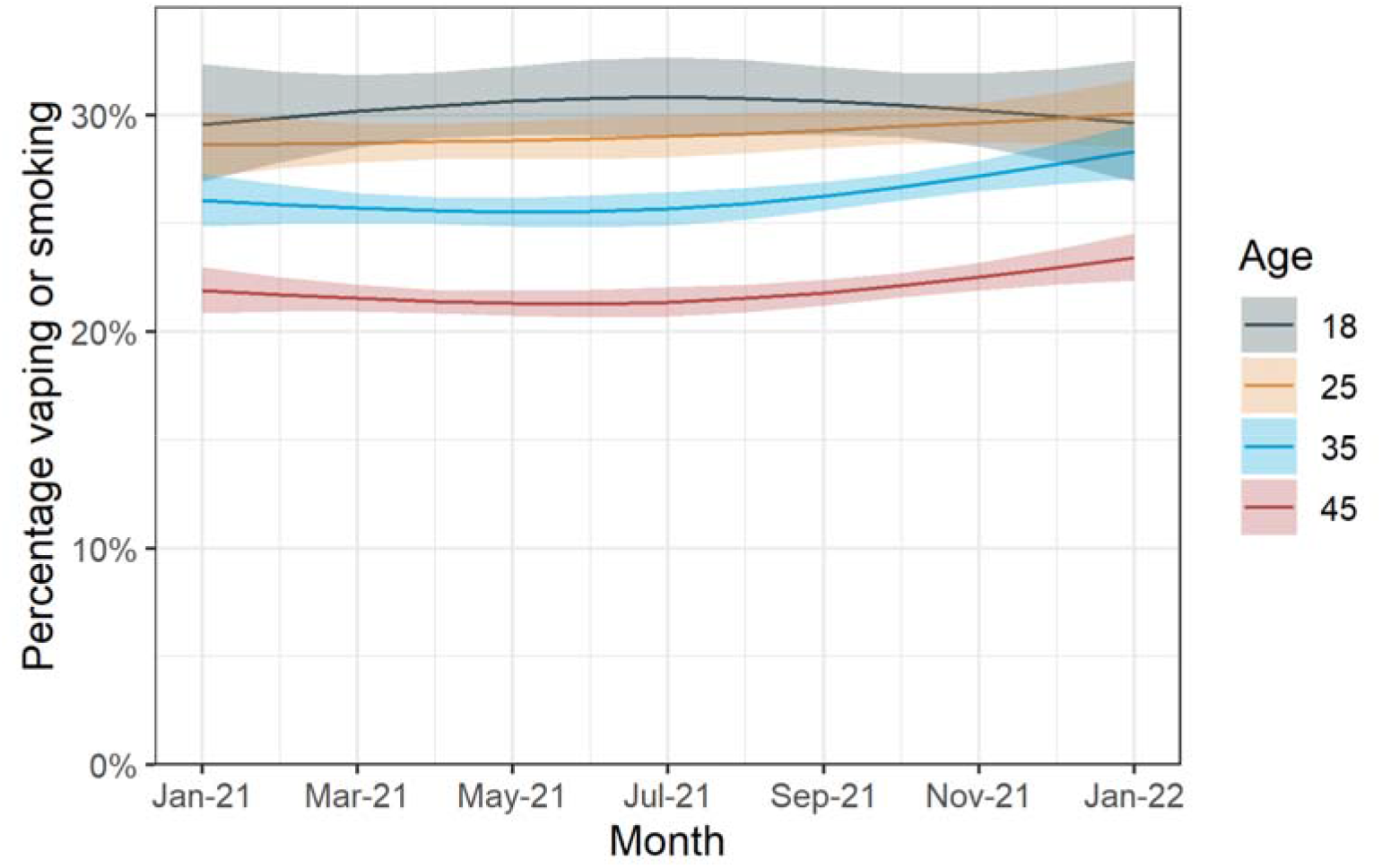
Prevalence of inhaled nicotine use, vaped or smoked, across ages in Great Britain from 2021 to January 2022. A total of 29,976 adults were surveyed (approximately 2,300 each month). Lines represent point estimates from logistic regression allowing an interaction between age and month, modelled non-linearly using restricted cubic splines. Shaded areas represent standard errors.

**Supplementary Figure 4.**
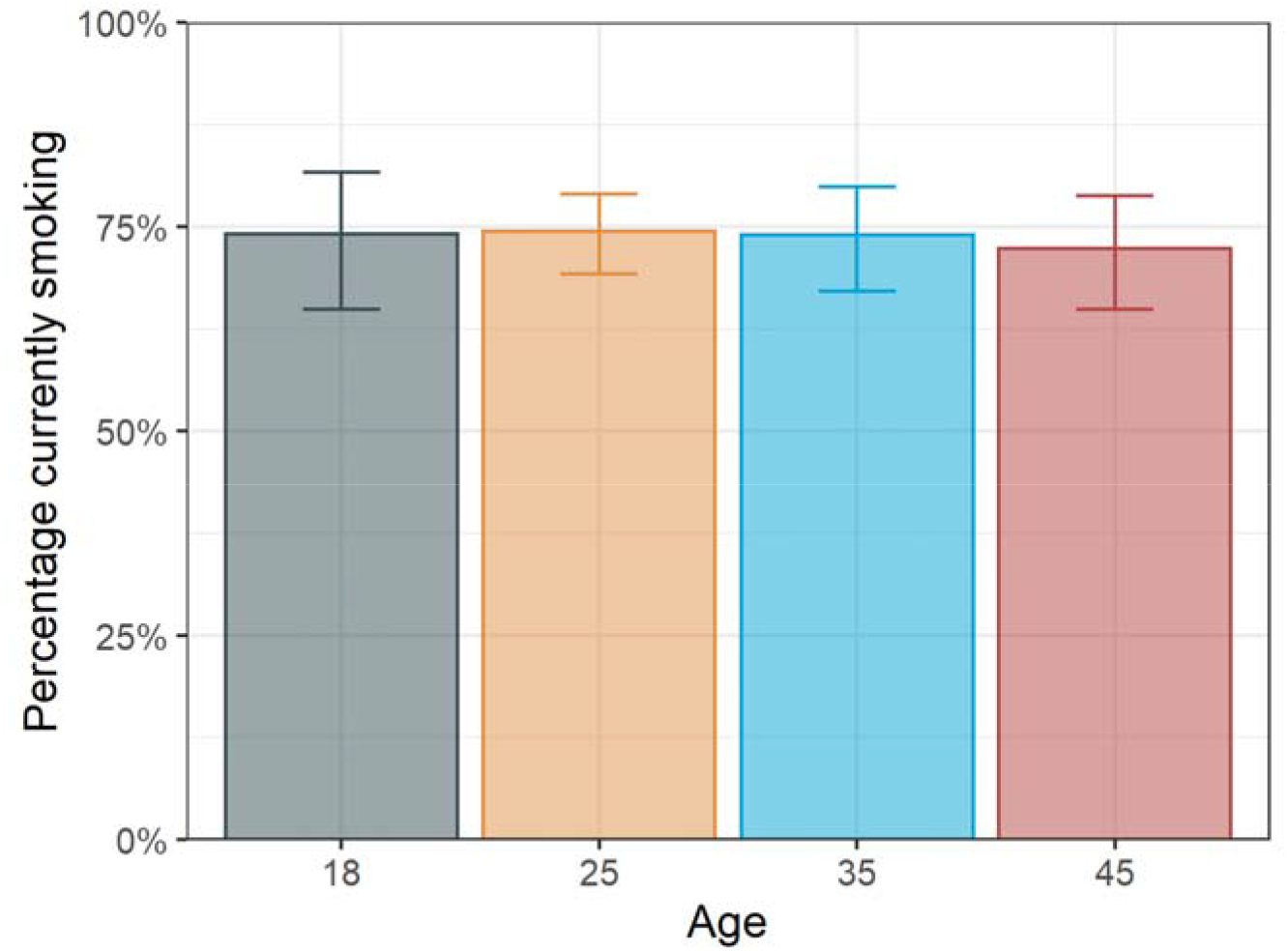
Percentage of disposable vapers who currently smoke across ages in Great Britain. Height of bars represent point estimates from logistic regression with age modelled non-linearly using restricted cubic splines. Error bars represent standard errors.

**Supplementary Figure 5.**
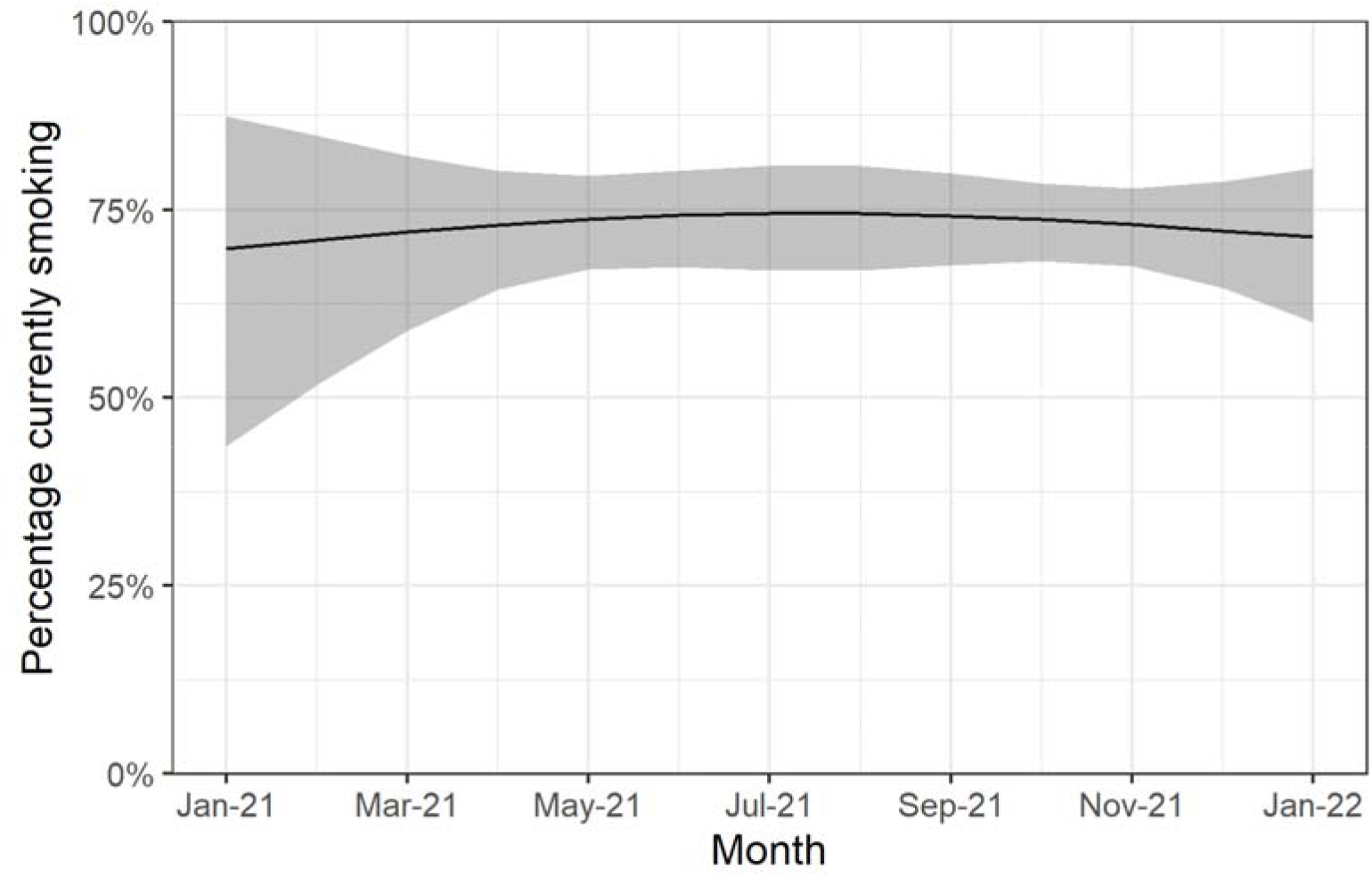
Percentage of disposable vapers who currently smoke across months from 2021 to January 2022 in Great Britain. Line represents point estimates from logistic regression with month modelled non-linearly using restricted cubic splines. Shaded bands represent standard errors.

